# A Timeline of Symptom Onset and Disease Progression in CLN3 Disease

**DOI:** 10.1101/2025.08.31.25334748

**Authors:** Ineka T. Whiteman, Anthony L. Cook, Erika F. Augustine, Aidan D. Bindoff, Alexandra M. Johnson, Heather L. Mason, Jonathan W. Mink, John R. Østergaard, Angela Schulz, Jennifer Vermilion, Amy Vierhile, Heather R. Adams

## Abstract

**Background:** CLN3 disease, or Juvenile Neuronal Ceroid Lipofuscinosis (JNCL), is a rare, genetic neurodegenerative condition, typically manifesting in the first decade of life and progressing in severity, with death typically occurring in early adulthood. Despite two decades of natural history research, a clear timeline of CLN3 disease symptom onset and progression remains poorly defined, limiting optimal patient management and therapeutic development. We conducted a literature review and analysed the natural history data to better understand the age of core symptom onset and chronological disease progression.

**Methods:** A literature review was undertaken using a pre-defined search strategy focused on CLN3 disease natural history studies, where age at onset for one or more core symptoms was reported in cohorts of ≥15 subjects. For each symptom, weighted mean age at onset and weighted standard deviation were calculated, with 95% confidence intervals derived from the weighted standard error. Symptom onset ages were compared using ANOVA.

**Results:** We identified nine natural history studies that met our pre-defined criteria. In total, 423 discrete patients aged between 4 and 39 years were reported. Thirteen core symptoms and a weighted average age at onset and weighted standard deviation were (in years): vision loss (6.1 ± 1.6, N=254), behavioural changes (8.5 ± 3.9, N=194), cognitive decline (9.3 ± 3.1, N=219), seizures (10.2 ± 3.0, N=243), sleep disturbance (11.0 ± 6.1, N=111), motor decline (11.0 ± 3.8, N=108), complete blindness (11.4 ± 3.6, N=171), speech and language impairment (12.7 ± 4.8, N=136), Parkinsonian gait (14.1 ± 2.5, N=111), cardiac manifestations (17.8 ± 4.4, N=45), loss of independent walking (19.5 ± 3.2, N=70), feeding difficulties requiring enteral feeding tube (22.0 ± 1.6, N=35), and death (22.4 ± 4.4, N=95).

**Conclusion:** This comprehensive summary of available natural history data illustrates mean age at onset of 13 core symptoms of CLN3 disease, and characterises a chronological timeline of disease progression. These results provide much-needed practical, anticipatory guidance to those involved in caring for individuals with CLN3 disease, and serve to highlight the critical importance of collecting globally standardised, quantifiable, longitudinal data for optimising patient management and advancing therapeutic approaches for CLN3 disease.

## Background

CLN3 (Batten) disease is a rare childhood-onset neurodegenerative disorder characterised by vision loss, progressive cognitive and motor decline, behavioural and mood symptoms, loss of speech and language, seizures, sleep disturbance, and feeding difficulties. In 80% of individuals, vision impairment is the first reported symptom (1, 2), occurring at around 5-6 years of age, often following an otherwise typical early childhood development. Symptoms emerge and worsen over an approximately 15–20-year period (3) and culminate in premature death during the twenties to thirties (2). To date, there are no approved disease-modifying therapies for CLN3 disease. Treatment remains limited to symptom management and palliation, with goals including mitigating the impact of symptoms and thereby optimising quality of life and reducing suffering.

CLN3 disease, historically called Juvenile Neuronal Ceroid Lipofuscinosis (JNCL), is one of 13 known neuronal ceroid lipofuscinosis (NCL) disorders, the most prevalent set of lysosomal storage disorders (4). Each of the 13 NCLs is classified according to the affected gene and age at onset, and although genetically distinct, all NCLs have a shared clinicopathology of accumulation of lysosomal storage material (5). CLN3 disease is caused by pathogenic variants in the *CLN3* gene, most commonly a 966 bp deletion (also referred to as “1-kb deletion”) of exons 7 and 8 on chromosome 16p11.2., predicted to result in a truncated protein. This deletion accounts for around 79% (6) of the affected alleles in CLN3 disease patients, typically giving rise to ‘classical’ CLN3 disease in terms of age at first symptom onset and general clinical disease progression. Approximately 74% of affected individuals are homozygous for the common deletion, and 22% are compound heterozygous, with other pathogenic variants present on the other allele. The remainder are compound heterozygous or homozygous for less common variants (7–9). Of the NCLs, CLN3 disease is one of the more common forms (9, 10) (11–13).

Owing to the heterogeneity and non-specificity of symptoms early in the disease course, along with the rarity and general lack of awareness of the disease, patients and their families often endure a ‘diagnostic odyssey’ during the years leading up to a Batten disease diagnosis (2, 14). This prolonged period of uncertainty, followed by a diagnosis of a severe, progressive, life-limiting condition and its ensuing relentless progression, has a significant emotional, physical and psychosocial impact on the affected individual and their family (15, 16).

In CLN3 disease, more than a dozen natural history studies conducted over the last two decades have characterised the salient and discrete clinical features of the condition, revealing a characteristic and predictable progression for the majority of affected individuals within the classical clinical phenotype and a linear relationship between disease severity and age (2, 14, 17–28).

Most individuals with CLN3 disease exhibit a ‘classic’ progression beginning with the onset of vision impairment, with cognitive impairment and behaviour and mood symptoms appearing simultaneously or during the following few years, followed by seizures, motor decline, and other symptoms. Other less commonly occurring clinical phenotypes associated with pathogenic variants on *CLN3* have been described, including a ‘protracted’ form with either a classical or delayed age of onset for vision loss and later onset of other symptoms (7, 29, 30), and a ‘vision loss’ only phenotype involving isolated retinal degeneration without progression to other symptoms (31–33).

Despite the numerous and ongoing natural history studies, to date, there has been no comprehensive summation of publicly available data that illustrates the full spectrum of CLN3 disease symptoms and the timeline of clinical progression beginning from symptom onset.

A clearer picture of disease trajectory may serve as a critical resource for parents, caregivers and clinicians, helping to reduce burdensome diagnostic delays and may provide the anticipatory guidance and care planning needed to optimise patient management and outcomes. In other severe diseases like Spinal Muscular Atrophy (SMA) and Duchenne Muscular Dystrophy (DMD), well-documented disease progression—derived from research, patient registries, and clinical experience—has helped caregivers recognise early symptoms such as hypotonia, difficulty feeding, and delayed motor milestones, allowing them to seek timely medical attention. For clinicians, understanding natural history provides a structured framework to enable earlier diagnosis, genetic testing, and initiation of disease-modifying treatments where available, alongside proactive interventions like respiratory support, physical therapy, and nutritional management tailored to each disease stage (34–37).

Similarly, a well-defined disease trajectory in CLN3 disease could facilitate earlier diagnosis and improve patient outcomes. We, therefore, undertook a review of the existing natural history literature on CLN3 disease to collate and critically evaluate available data on key symptoms and their age of onset.

## Methods

We reviewed the current natural history literature on CLN3 disease using the PubMed database (accession date: June 2024). Search terms were ‘CLN3 phenotype’, ‘juvenile batten phenotype’ and ‘CLN3 symptom’, focused on English-language publications but without using other filters. Studies were included if they reported data from cohorts of 15 or more individuals with genetically confirmed CLN3 disease, where ages of symptom onset were provided via clinical records or caregiver interviews. We included cohorts of n=15 or larger, as these studies were considered sufficiently robust as sources of meaningful data. Individual reports and small case series were excluded. To minimise the possibility of individual patients being reported across multiple studies, we excluded studies that investigated the same or overlapping cohorts, as determined by the study site, except where studies investigated different symptoms in the same cohorts. For example, studies conducted by Cialone et al. (2), Augustine et al. (27) and Masten et al. (26) reported on many of the same research participants (though sample sizes vary across these studies) enrolled in the University of Rochester Batten Center (URBC, Rochester, NY) natural history study. Given that Cialone et al. (2) assessed the greatest number of symptoms, this study was retained for the statistical analysis, while the studies by Augustine et al. (27) and Masten et al. (26) were excluded. In contrast, since Nielsen and Østergaard (38) and Østergaard (18) assessed different symptoms from the same cohort (recruited from Aarhus University Hospital, Denmark), both studies were included in our statistical analysis.

We extracted data relating to the age of onset for each symptom reported, including mean and standard deviation (SD), median, range, and interquartile range (IQR), depending on data available (Table 1). We initially calculated the weighted mean age for the first onset of each symptom, where each study was weighted according to the number of participants, regardless of whether SD, range or IQR was reported, which yielded a simple *weighted mean age* of onset and overall largest pooled sample sizes (Table 1).

**Table 1.**
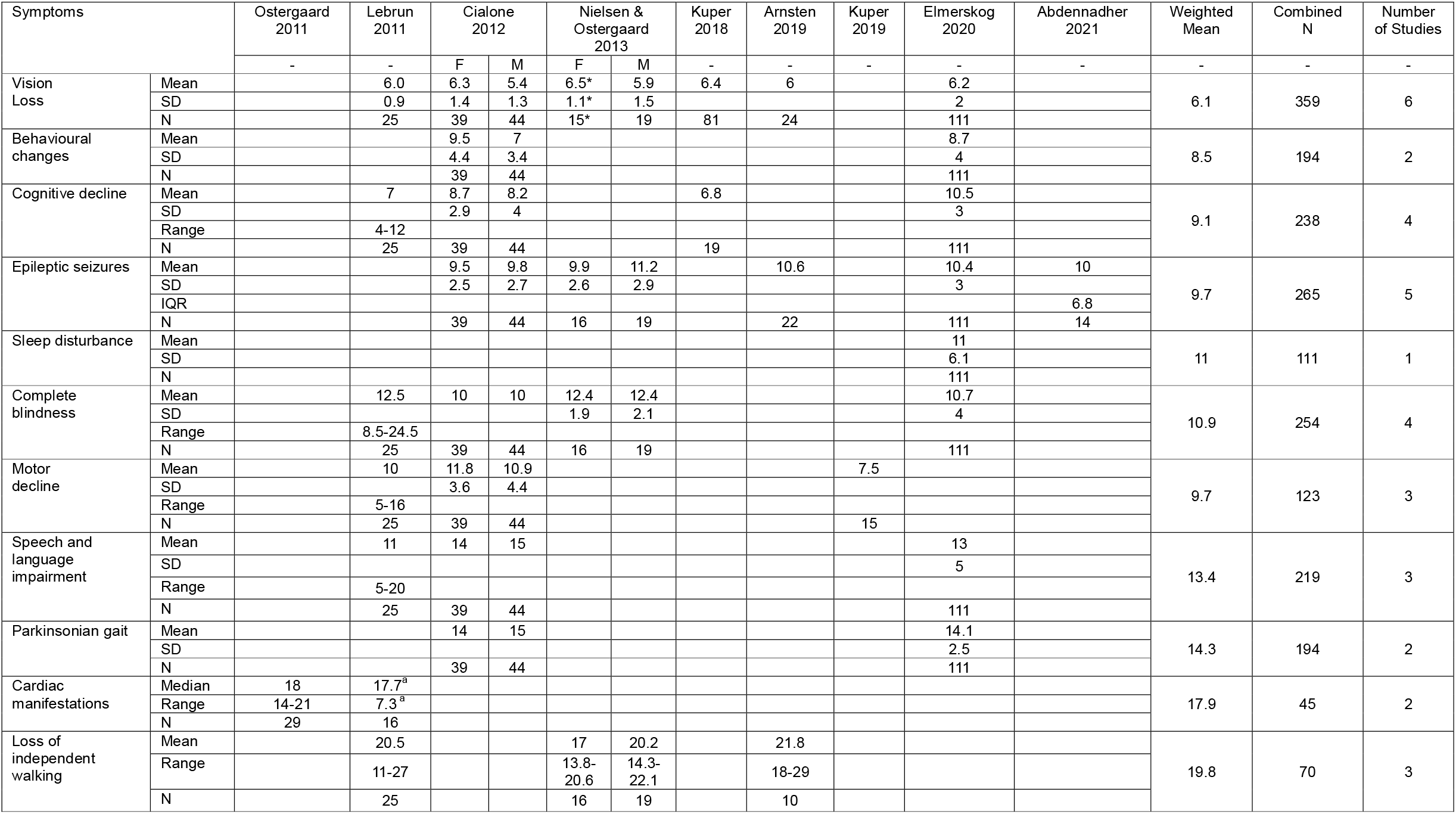
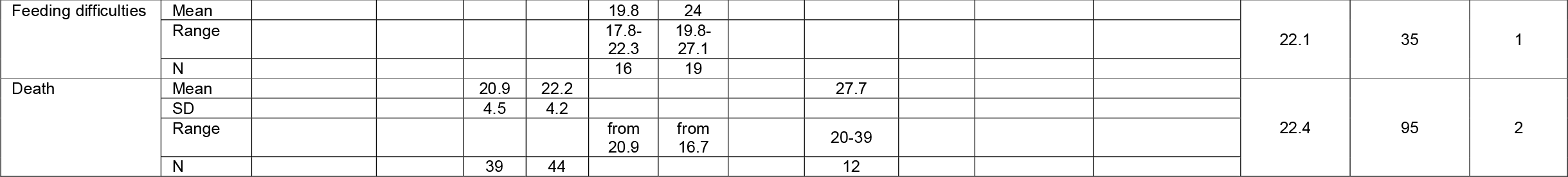
Age at onset for core symptoms of CLN3 disease. For each symptom, mean age at onset and SDs or min/max ranges (depending on available data) are presented from nine discrete natural history studies (see Refs). Calculated weighted means, total number of studies and total combined N for each symptom are also included in the columns on the right. Abbreviations: F, female; M, male; IQR, interquartile range; SD, standard deviation. ^*^(J. Ostergaard, MD, email communication, 02 April 2025: pertaining to mean [SD] age of onset of vision loss for n=15 females in sample; review of data not reported in published manuscript)

Given the rarity of these important natural history data points and the varying analysis methods used across studies, rather than excluding valuable data, we transformed reported datasets in order to present pooled data and robustly compare across symptoms. To do this, we used the *Method for Unknown Non-Normal Distributions* formula (39) to transform median/mean and range reported in four studies (18-20, 38) to mean and SD. Similarly, the IQR reported by Abdennadher et al. (17) was used to estimate SD using the formula SD = IQR/1.35. Data from these transformation steps are presented in Supplementary Table 1. These transformed data were combined with reported means and SDs and used to calculate weighted mean, weighted SD and combined N derived from all available studies for all core clinical symptoms of CLN3 disease (Supplementary Table 2), which were further analysed by a Brown-Forsythe ANOVA test for equality of means (as groups had unequal variance). In this analysis, the weighted mean age of onset of each symptom was compared to all other symptoms in the dataset using Games-Howell’s multiple comparisons test (GraphPad Prizm version 10.3.1), where an adjusted P < 0.05 was considered significant (Figure 1A-B).

**Figure 1.**
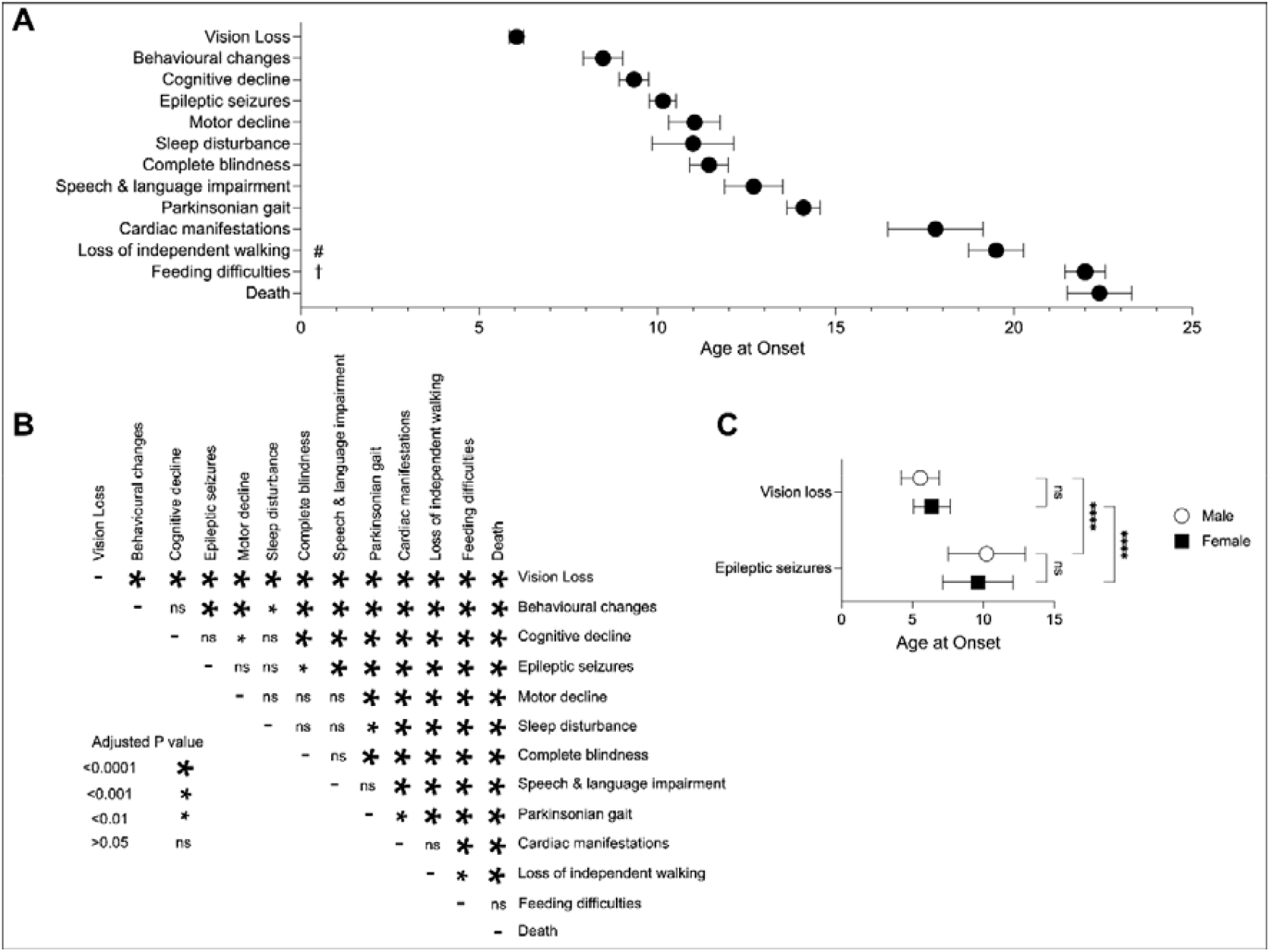
Timeline of disease progression in classical CLN3 disease. **(A)** Timeline of weighted mean age and 95% confidence interval for the initial onset of the core clinical symptoms in CLN3 disease. # Unable to walk without assistance or daily use of wheelchair; † Requiring enteral feeding tube. (**B**) Results of ANOVA test for all pairwise comparisons of weighted mean age at onset for core clinical symptoms of CLN3 disease. Size of the asterisk indicates adjusted P value as described in panel key; for each symptom, N ranges from 35–254 participants, derived from 1-4 studies. (**C**) Weighted mean ages ± weighted standard deviation of initial onset for vision loss and epileptic seizures were compared for males (N=63; open circles) and females (N=54-55; black squares) using a 2-way ANOVA test; ^****^ = P < 0.0001, ns = P > 0.05.

Two studies reported sex-specific data for several symptoms (2, 38), enabling the comparison between sexes for age of onset for vision loss and epileptic seizures. Sex-specific weighted means and weighted SD were calculated from these studies and compared using a 2-way ANOVA test and Šídák’s multiple comparisons test (GraphPad Prizm), where an adjusted P < 0.05 was considered significant (Figure 1C and Supp. Table 2).

## Results

The literature search yielded 548 publications, of which 153 were duplicates. The remaining 395 publications included nine natural history studies that evaluated one or more clinical symptoms of CLN3 disease in different patient cohorts. In total, 423 affected individuals were included in the present analysis, with a reported age range between 4 and 39 years.

### Age at Diagnosis

Similar ages at diagnosis were reported by Kuper et al. (2019) at 7 years (range 5-12) (22) and Arntsen at 7.3 years (range 3-13 years) (20). In the JNCL and Education Project Survey (n=111), parents reported that the average age at which they sensed a concern for their child was 5.6 years, with visual impairment reported at 6.2 years and a diagnosis at 8.0 years (23). Of note, genetic diagnosis of CLN3 disease was not independently confirmed, but many were presumed to have this diagnosis based on parent-reported information regarding age of onset, symptom presentation, and/or family history.

#### Sex differences in age at diagnosis

Reported in one study, the average age at diagnosis was 7.9 years (± 1.2) for females and 7.0 years (± 1.1) for males (38).

### Onset of Vision Loss and Complete Blindness

Visual impairment is typically the first reported symptom of CLN3 disease. Vision loss in CLN3 disease is characterised by an initial loss of central vision progressing rapidly to total blindness over approximately five years. Early characteristics of vision loss in CLN3 disease overlap with those of other inherited diseases of the eye (e.g., retinitis pigmentosa, Stargardt disease) and other progressive cone-rod dystrophies, which can contribute to initial misdiagnosis and a lengthening of the diagnostic odyssey (40–42). Six of the nine studies meeting inclusion criteria reported the age of onset of vision loss, with a combined average age of 6.1 years (N = 359; Table 1). In a longitudinal study by Lebrun et al. (N = 25; 11 male and 14 female), all children shared the same variant in the *CLN3* gene, a homozygous 1 kb deletion. Data were collected over a mean period of 18.4 years (± 6.2 years). Data were collected using an established clinical scoring system (24). The first clinical sign in 17 patients was vision deterioration at age 5.4 years (± 1.4)(19). Data from all 25 subjects revealed a mean age at onset of vision loss to be 6.0 years (± 0.9 years; A. Schulz, MD. Email communication. 2025 Jul 04). From four studies, we calculated a weighted mean of 6.1 ± 1.6 years (N = 254), which was a significantly younger age than all other symptoms (Figure 1A-B and Supp. Table 2).

The age at which vision was lost completely was reported in four studies (2, 19, 23, 38). The average age at complete loss of vision was 10.9 years (N = 254) (Table 1), though it should be noted that ‘complete’ vision loss was not consistently defined across the studies. In our pooled analysis of three studies (19, 23, 38), complete loss of vision occurred by 11.4 years ± 3.6 (N = 171) (Figure 1A-B and Supp. Table 2). Cialone et al. and Elmerskog et al. both report complete vision loss at around ten years of age (2, 23), whereas Lebrun et al., as well as Nielsen and Østergaard, report around 12.5 years (19, 38).

#### Sex differences in onset of vision loss

Nielsen & Østergaard (38) report visual impairment as the first symptom in 34 of 35 affected individuals (97%), and that there was no difference in age of vision loss between sexes, with an average age of onset of 6.4 years (± 1.1) for females and 5.9 years (± 1.1) for males (Table 1; J Ostergaard, MD. Email communication, 2025 April 02). In contrast, Cialone et al. (2) found that the onset of vision loss began one year later in females (6.2 ± 1.4 years) than in males (5.2 ± 1.0 years) (Table 1), but this difference was not identified in our pooled analysis (P = 0.11 [N = 54 female and 63 male (Fig 1C and Supp. Table 2].

### Behavioural and Psychiatric Symptom Onset

Behavioural and psychiatric symptoms in CLN3 disease are variable in their expression and severity and may not occur in all affected individuals. Children may experience an increase in challenging behaviours, including physical and verbal aggression, rage episodes, noncompliance, and defiance (15, 43–45). Aggression may be directed at others or may involve self-injurious behaviours. A commonly described concern is perseverative behaviour characterised by repetitive thoughts and persistent focus on particular ideas or preferred activities; these may or may not be accompanied by agitation when perseverative ideas or activities cannot be carried out to the child’s satisfaction. Anxiety is also a common mood symptom. Though less common, auditory and/or visual hallucinations have also been reported in some individuals (15, 46, 47).

While vision loss is the most common first symptom, behavioural impairment was also reported as an early symptom in two studies (2, 23). Lebrun et al. reported that “behavioural problems” were the first symptom in five children (20%); the authors did not describe the nature of those problems or how they were assessed, but reported that they became apparent at a comparable age of onset to vision loss (5.8 ± 1.1 years). In Cialone et al.’s study of 83 children (39 female, 44 male), parents reported “behavioural difficulties” (p. 551) as an initial symptom in 11 (13%) individuals. (2). In our pooled analysis, the average age of onset of behavioural change was 8.5 ± 3.9 years (N = 194) (Figure 1A and Supp Table 2).

#### Sex differences in onset of behavioural and psychiatric symptoms

Cialone et al. reported that, on average, behavioural symptoms begin later in females (9.5 ± 4.4 years) than in males (7.0 ± 3.4 years; p = <0.05) (2) (Table 1).

### Cognitive Decline Onset

Because of early vision loss in most children with CLN3 disease, formal assessment of cognition has focused on verbally mediated skills. Cognitive difficulties in CLN3 disease include impairments in attention, new learning and recall, verbal fluency, and verbal intellectual ability (IQ) (21, 23, 48, 49). The onset of cognitive difficulties has been reported in four studies, which found that the average age when cognitive changes become apparent is 9.1 years (N = 238). In an educational setting, initial difficulties at school are commonly attributed to visual deterioration. However, when performance on intelligence (IQ) tests was compared to patients with Stargardt disease who have isolated vision loss, it was clear that for children with CLN3 disease, the cognitive impairments were already present around the time of diagnosis (21). Most investigations have described early cognitive changes as representing ‘decline’ due to numerical decreases in age-adjusted scores in relation to same-age peers. However, this interpretation overlooks a limitation of these age normative-referenced scores, which decrease when there is a slowing of cognitive growth compared to same-age peers (50, 51). In fact, in the early years of cognitive change in children with CLN3 disease, affected children may not experience a loss of previously attained skills. Rather, there may be a pattern of initial slowing of cognitive development, followed by a plateau in the acquisition of new skills, then followed by loss of previously attained abilities (i.e., cognitive decline) (23). This is a common pattern reported amongst children with inherited neurodegenerative diseases and, in concert with behavioural and psychiatric/mood symptoms, can be considered as childhood-onset dementia (52).

The onset of cognitive changes was reported in four studies (2, 19, 21, 23), allowing pooled analysis, which revealed an average age of onset of 9.1 years (N = 238) (Table 1). Three studies reporting mean and SD could be combined (2, 19, 23), revealing the onset of cognitive changes by 9.3 ± 3.1 years (N = 219) (Figure 1A and Supp Table 2). As noted above, changes in cognition are characterised by a deceleration or plateau in cognitive growth before cognitive decline is observed (23). The timing of the neurocognitive changes in classical CLN3 disease was described by Kuper et al. 2018 (21). Their literature-derived patient descriptions and a retrospective single⍰centre Dutch cohort showed an onset of visual impairment at 6.4 years (range 4-9 years) (N = 81) with paralleled cognitive changes beginning at a mean age of 6.8 years (range 2-13) (N = 19) (21). Lebrun noted the onset of cognitive changes slightly later, at a mean age of 7 years (range 4-13 years) (19). In protracted CLN3 disease, cognition may remain normal until adulthood (21).

#### Sex differences in onset of cognitive impairments

There appear to be no significant differences between females and males in the onset of cognitive impairments (8.69 vs 8.22 years) (2).

### Seizure Onset

The most common seizure type in individuals with CLN3 disease is generalised tonic-clonic seizures (2, 27), reported in one paper as occurring in up to 78% of affected individuals (27). Other seizure types include complex partial, absence, myoclonic, atonic and focal seizures. Fifty per cent of affected individuals experienced a single seizure type that mildly worsened with age. Except for myoclonic seizures, seizures occurred less than once every three months, which were usually managed with one or two anti-seizure medications (27).

Five studies reported the age of seizure onset, with the combined mean age of onset of 9.7 years (N = 265) (Table 1) (2, 17, 20, 23, 38). Four studies could be combined for our pooled analysis (2, 17, 23, 38), which revealed an average age of onset for seizures of 10.2 ± 3.0 (N = 243) years (Figure 1A and Supp Table 2).

#### Sex differences in seizure onset

Nielsen and Østergaard compared sex differences in seizure onset in a Danish population of 19 males and 16 females with genetically confirmed CLN3 disease (38), reporting no differences in seizure frequency or anti-epilepsy drug (AED) use but that the first epileptic seizure occurred earlier in females than males (9.9 years ± 2.6 vs 11.2 years; ± 2.9 years) (38) (Table 1). In contrast, no significant differences between the age at onset of seizures between females and males were reported by Cialone et al. (9.5 vs 9.9 years, respectively) (2) (Table 1). Our combined analysis of these two datasets showed no difference in age of onset for seizures between females and males (P = 0.38 [N = 55 females and 63 males]; Figure 1C and Supp. Table 2).

### Sleep Disturbance

Children with CLN3 disease may experience various sleep problems, including sleep onset and maintenance difficulties, fragmented and irregular sleep, reduced total sleep time and sleep efficiency, and increased daytime sleepiness. Sleep problems may worsen with advancing age and/or disease severity (53, 54). However, small, clinically heterogeneous samples and variable methods limit our current understanding of the scope and severity of sleep disturbances in this population. Nonetheless, sleep disturbances are most commonly reported as developing early in the second decade of life (20, 23). Ages of onset for sleep disturbances were only reported in one of the nine studies included in our analysis (23), occurring at 11.0 ± 6.1 years (N = 111) (Figure 1A and Table 1). No known studies have examined sex differences in sleep function and/or disordered sleep in individuals with CLN3 disease.

### Motor Dysfunction Onset

The onset of mobility issues was reported in three studies (2, 19, 22) with a mean average age for onset of motor decline of 9.7 years (N = 123) (Figure 1A and Table 1). We combined mean and SD data from two studies (2, 19), determining an age of onset of 11.0 ± 3.8 years (N = 108) (Figure 1A and Supp. Table 2); we note the lower average age of onset in this analysis is due to the notably lower age of onset reported in Kuper et al (22) (7.5 years) but which was not able to be transformed.

The 6-Minute Walk Test (6MWT) was employed by Kuper et al. (2019) to evaluate motor function twice per year in 15 children (65 tests) with genetically confirmed CLN3 disease (22). In this standardised assessment of functional exercise capacity, lower Z-scores reflect greater impairment. Comparisons were made between children with CLN3 disease and two control cohorts: children with visual impairment without cognitive deficit (n = 14), and children with visual and non-progressive neurological impairment (n = 12). The mean 6MWT Z-scores for the CLN3 children were impaired shortly after diagnosis of −3.6 (n = 3) and −4.7 (n = 5) at 7 and 8 years, respectively. In contrast to the control groups, their scores declined significantly with age, suggesting motor impairment is not necessarily explained by potentially confounding visual or cognitive impairments, but intrinsic to the CLN3 disease process itself.

#### Sex differences in onset of motor declin

Cialone et al. reported there were no significant differences between females and males in the onset of motor decline (11.8 vs 10.9 years) (2) (Table 1).

### Loss of Independent Walking

The age at which individuals were unable to walk without assistance or required daily wheelchair use was reported as a mean and range in three publications (19, 20, 38). The age of onset for loss of independent walking varied among studies. Arntsen et al. reported the mean age at which patients lose the ability to walk independently to be 21.8 years (range 18 −29) (20), whereas Lebrun et al. reported a much wider range (11 – 27 years) but a similar average age of onset of 20.5 years (19) (Table 1). The weighted mean age for loss of independent walking from these three studies was 19.8 years (N=70; Table 1). After transformation, data from these studies were used to calculate weighted mean, weighted SD and combined N, revealing a weighted mean age of onset of 19.5 ± 3.2 years (N = 70) (Fig. 1A, Supp. Table 1).

#### Sex differences in loss of independent walking

A study conducted by Nielsen and Østergaard found that wheelchair dependency was significantly earlier in females than males (17.0 vs 20.2 years, respectively) (38) (Table 1). Cialone et al. also described a sex-based difference in loss of functional independence in CLN3 disease, with females losing independence approximately one year earlier than males from the age at initial symptom onset (12 vs 13 years) (2) (Table 1).

### Speech and Language Impairment

Children with CLN3 disease develop speech impairment early in the disease course, with dysarthria often manifesting in a characteristic neurogenic stuttering (55). Over time, speech clarity deteriorates due to motor speech impairment, as does verbal output. Individuals with CLN3 disease may demonstrate stronger expressive language than receptive language skills, based on parent report (55). Speech impairment, reported in three studies, began, on average, at 13.4 years of age (N = 219) (Table 1) but had a very wide range, occurring as young as five years and as late as 20 years of age. Word-finding (n = 83) and dysfluency (n = 47) are described as the most frequent specific issues (23, 55). Combining data from two studies (19, 23) resulted in an average age of onset of 12.7 ± 4.8 years (N = 136) (Figure 1A and Supp. Table 2). The mean age for unintelligible speech is around 20.3 years (23). No known studies have examined sex differences in speech and language impairment in individuals with CLN3 disease.

### Feeding Difficulties

Only one study reported feeding difficulties requiring an enteral feeding tube at an average of 22.1 years (range 18-27; N=35). Mean and range were transformed, revealing a weighted average age of onset of feeding difficulties and estimated SD to be 22.0 ± 1.6 years (N = 35) (Fig. 1A, Supp. Table 1 and 2). Feeding complications requiring a gastric tube occurred more than four years earlier in females than males (19.8 vs 24.0 years, respectively) (38) (Table 1).

### Cardiac Manifestations

Late in CLN3 disease, affected individuals may develop cardiac issues, including sinus node dysfunction, bradycardia and other conduction abnormalities (1, 56). The mean age of CLN3 patients with pathologic cardiac findings was 17.7 ± 7.3 years (mean ± SD, range 11–34 years) (19). In the current analysis, cardiac symptoms of CLN3 disease were analysed from this and one other study (18, 19), the latter also with a wide range for age of onset, between 14 and 21 years of age (Table 1). The combined mean age of onset was 17.9 years (N=45) (Table 1). Analysis of transformed data revealed a weighted average age of onset of cardiac manifestations and estimated SD to be 17.8 ± 4.4 years (N = 45) (Fig. 1A, Supp. Table 1 and 2).

### Premature Death

The life expectancy of people with classic CLN3 disease is twenty to thirty years (18, 27), with a range of 20-39 years (2, 20). Causes of death are primarily attributed to congestive heart failure, infection, status epilepticus, and episodes suggestive of autonomic dysfunction (18, 20, 38). Two studies reported the mean age of death with a weighted mean of 22.4 years (N=95; Table 1). Combining data following a transformation step for Arnsten et al. (Supp. Table 1) revealed a mean age of onset and estimated SD of 22.4 ± 4.4 years (N=95; Figure 1A, Supp. Tables 1 and 2). A third study, Nielsen and Østergaard (38), reported death to occur from 20.9 years in females (n=16) and from 16.7 years in males (n=19) (Table 1); however, the mean age of death was not reported, excluding this study from the pooled analysis.

#### Sex differences in age of death

Cialone et al. (2) reported that death was significantly earlier in females (20.9 ± 4.5 years) compared to males (22.2 ± 4.2 years; Table 1). Nonetheless, their analysis was restricted to individuals < 30 years of age.

### Combined Natural History Analysis

We next compared the ages of onset for these 13 core symptoms, using our pooled dataset derived from published and transformed data, by performing a 1-way ANOVA (Figure 1B). This analysis revealed that vision loss occurred at disease onset at 6 years of age, significantly earlier than all other symptoms. This was followed by a period of relatively rapid progression, in which eight symptoms occurred between approximately 8.5 and 14 years of age. Within this group, some symptoms occurred at significantly different ages (e.g. behavioural changes at age 8.5, compared to complete blindness at age 11.4, compared to Parkinsonian gait at age 14.1). However, our analysis also revealed substantial overlap for age at onset for some symptoms. For example, age at onset for motor decline, sleep disturbance, complete blindness, and speech and language impairment were not significantly different, and all four of these symptoms typically occurred within an approximately 18-month period between ages of 11 and 12.7 years. At late stages of the disease course, cardiac manifestations and loss of independent walking occurred at similar ages (18 or 19.5 years), but earlier than feeding difficulties and death, which occurred together at 22 years.

### Discussion

We have presented for the first time a cohesive and comprehensive summary of published natural history data characterising the typical onset and pattern of progression of 13 core clinical symptoms of CLN3 disease. These data, collated from nine discrete studies and cohorts, provide the largest grouped data available in the literature for CLN3 disease and should reduce bias present in smaller cohorts. The first symptom of CLN3 disease is typically vision loss, which is consistently reported across the literature with an onset at around six years of age, although there is a range from 3 to 7 years old. Additionally, variability is seen in the age at onset of all symptoms of CLN3 disease reported here. It is unlikely that phenotypic heterogeneity can be explained by CLN3 genotype alone, but future research is needed to explore potential epigenetic and/or other contributions to variability in symptom onset and expression. More recently, with the growing understanding and clinical experience of CLN3 disease, additional symptoms of potential core clinical relevance have been recognised. Bowel dysfunction, including faecal incontinence and constipation, often becomes prominent as the disease advances, leading to complications such as malnutrition, abdominal distension, abdominal pain, and diminished quality of life (1, 57).

Furthermore, a behaviour-related symptom has been increasingly observed in later stages: fearful, anxious, and aggressive behaviours. While these features are only qualitatively described in some of the natural history studies reviewed, they are consistently reported by families as distressing and burdensome, particularly in older patients, and are supported by clinical case reports (15, 46, 47, 58). Further investigation into the aetiology and management of these behaviours is warranted, given their impact on quality of life, and may be considered for explicit inclusion in clinical rating scales and future natural history research.

We acknowledge several limitations in comparing data amongst individual studies and cohorts across multiple and international sites, as well as in our statistical approach. First, there is variability in data collection methods, extraction and reliability, with some studies collecting prospective longitudinal data using standardised clinical assessments and outcome measures (17–20, 22). In contrast, others are retrospective, often collected through caregiver reports unverified by a clinical researcher or other independent source (2, 21, 23, 38). Second, there is wide variation in patient cohort size between studies, per analysed symptom, or both, and some symptoms (e.g. sleep disturbance, Parkinsonian gait, feeding difficulties) are reported only by one study included in our pooled dataset; further data on the age of onset for those symptoms reported from fewer participants might affect findings from the present analysis despite attempts to mitigate this limitation by calculating weighted means and SD. Last, we analysed data reported in differing formats, necessitating us to estimate means and standard deviations for some or all symptoms reported in some studies (see Supplementary Table 1). While we used an approach that accounts for skewed distributions (39), consistent reporting of mean and SD across future studies would maximise the value of each study. This is particularly important for CLN3 disease and other forms of neuronal ceroid lipofuscinoses, given the rarity of the condition. Nonetheless, we are encouraged by results that are qualitatively consistent between studies, particularly in three key symptoms where a consistent mean age of onset and low variance were observed (vision loss, seizures and complete blindness).

The present study is truly patient-centric in its conception, borne out of the expressed need for such a resource by affected families and healthcare communities. To date, a resource of this nature has been notably lacking, and we anticipate it will be of great practical value to affected families, multidisciplinary healthcare teams, allied health professionals, support services, insurance providers and others. One of the key benefits of a holistic understanding of CLN3 symptomatology and progression is to provide anticipatory guidance, personalised care, and support of future planning, thereby ensuring the increasing medical, psychosocial, financial, and healthcare insurance needs of patients and their families are adequately met.

In addition to the benefits to families and providers, noted above, the advantages of a holistic understanding of CLN3 symptomatology and progression are multi-fold. Although these data will not be suitable for regulatory agencies, our data may also identify important knowledge gaps, thereby informing the design and development of current and future CLN3 natural history studies. A cohesive understanding of CLN3 disease natural history may also help guide clinical trial design, informing the selection of appropriate clinical endpoints relative to the age and stage of the CLN3 disease trajectory. Given the vital role of robust natural history data in therapeutic development for Batten disease (59), this study highlights the ongoing need to collect standardised, precise datasets on quantifiable measures of CLN3 symptoms and disease progression across sites and studies globally.

### Conclusion

The current lack of comprehensive and standardised natural history data for CLN3 disease represents a significant gap in the field. This analysis delineates the typical age at onset for core symptoms and the characteristic trajectory of classical CLN3 disease progression. Future optimisation of clinical management, implementation of evidence-based anticipatory care, informed trial design, and – critically – the establishment of benchmarks for evaluating the efficacy of emerging therapies all depend on the systematic collection of high-quality, quantitative, and standardised clinical data in CLN3 disease, ultimately ensuring that advances in research translate into meaningful improvements for patients and their families.

## Data Availability

All data produced in the present work are contained in the manuscript

## Declarations

### Ethics Approval And Consent To Participate

‘Not applicable’

### Consent For Publication

‘Not applicable’

### Availability Of Data And Materials

The datasets used and/or analysed during the current study are available from the corresponding author upon reasonable request.

### Competing Interests (From Each Author)

Each author has completed the ICJME declaration forms, none of which indicated any competing interests that may have influenced the validity of this research. Copies of the completed ICJME forms are available upon reasonable request.

### Author’s Contributions

Each author has approved the submitted version, along with supporting documentation, and has agreed both to be personally accountable for the author’s own contributions and to ensure that questions related to the accuracy or integrity of any part of the work, even ones in which the author was not personally involved, are appropriately investigated, resolved, and the resolution documented in the literature.

## Acknowledgements

This project was inspired by the families and healthcare community seeking to better understand and plan for the time course of CLN3 disease. The authors wish to thank the Beyond Batten Disease Foundation, the Batten Disease Support, Research and Advocacy (BDSRA) Foundation and BDSRA Australia for providing funding support for this project.

