## Supplementary Materials for "A Timeline of Symptom Onset and Disease Progression in CLN3 Disease"

### Supplementary Table 1. Estimated Data from Transformation of Reported Datasets.

| Symptoms | | | Ostergaard 2011[a] | | Lebrun 2011[b] | | Nielsen & Ostergaard 2013[b] | | | | Arnsten 2019[b] | | Abdennadher 2021[c] | |
|---|---|---|---|---|---|---|---|---|---|---|---|---|---|---|
| | | | - | | - | | Female | | Male | | - | | - | |
| | | | Reported | Estimated | Reported | Estimated | Reported | Estimated | Reported | Estimated | Reported | Estimated | Reported | Estimated |
| Cognitive decline | Mean | | | | 7 | 7.24 | | | | | | | | |
| | SD | | | | | 2.05 | | | | | | | | |
| | Range | | | | 4-12 | | | | | | | | | |
| | N | | | | 25 | | | | | | | | | |
| Epileptic seizures | Mean | | | | | | 9.9 | | 11.2 | | 10.6 | | 10 | |
| | SD | | | | | | 2.6 | | 2.9 | | | | | 5.04 |
| | IQR | | | | | | | | | | | | 6.8 | |
| | N | | | | | | 16 | | 19 | | 22 | | 14 | |
| Complete blindness | Mean | | | | 12.5 | 13.34 | 12.4 | | 12.4 | | | | | |
| | SD | | | | | 3.61 | 1.9 | | 2.1 | | | | | |
| | Range | | | | 8.5-24.5 | | | | | | | | | |
| | N | | | | 25 | | 16 | | 19 | | | | | |
| Motor decline | Mean | | | | 10 | 10.11 | | | | | | | | |
| | SD | | | | | 3.01 | | | | | | | | |
| | Range | | | | 5-16 | | | | | | | | | |
| | N | | | | 25 | | | | | | | | | |
| Speech and language impairment | Mean | | | | 11 | 11.37 | | | | | | | | |
| | SD | | | | | 4.08 | | | | | | | | |
| | Range | | | | 5-20 | | | | | | | | | |
| | N | | | | 25 | | | | | | | | | |
| Cardiac manifestations | Median | | 18 | | | | | | | | | | | |
| | Mean | | | 17.86 | 17.7 | | | | | | | | | |
| | Range | | 14-21 | | | | | | | | | | | |
| | SD | | | 1.77 | 7.3 | | | | | | | | | |
| | N | | 29 | | 16 | | | | | | | | | |
| Loss of independent walking | Mean | | | | 20.5 | 20.03 | 17 | 17 | 20.2 | | 21.8 | 22.2 | | |
| | Range | | | | 11-27 | | 13.8-20.6 | | 14.3-22.1 | | 18-29 | | | |
| | SD | | | | | 4.51 | | 1.87 | | 2.26 | | 3.22 | | |
| | N | | | | 25 | | 16 | | 19 | | 10 | | | |
| Feeding difficulties | Mean | | | | | | 19.8 | 19.8 | 24 | 23.81 | | | | |
| | Range | | | | | | 17.8-22.3 | | 19.8-27.1 | | | | | |
| | SD | | | | | | | 1.23 | | 1.99 | | | | |
| | N | | | | | | 16 | | 19 | | | | | |
| Death | Mean | | | | | | | | | | 27.7 | 27.92 | | |
| | SD | | | | | | | | | | | 5.47 | | |
| | Range | | | | | | from 20.9 | | from 16.7 | | 20-39 | | | |
| | N | | | | | | | | | | 12 | | | |

[a] Mean and SD were estimated using the formula described by Cai et al.
[b] We assumed a normal distribution and therefore that the reported means were an approximation of the median. The reported mean (approximated median) was then used with the reported range and N to estimate mean and SD using the formula described by Cai et al.
[c] SD was estimated by dividing the IQR by 1.35
**SD**, standard deviation; **IQR**, interquartile range; **M**, male; **F**, female

### Supplementary Table 2: Combined Dataset for Core Symptoms of CLN3 Disease

| | **Studies with Mean (+/- SD)**<br>**Studies with transformed Mean (+/- SD)** | | | | **Sex Differences** | | | | | | |
|---|---|---|---|---|---|---|---|---|---|---|---|
| | *Relates to Figure 1A-B* | | | | *Relates to Figure 1C* | | | | | | |
| Symptom | Weighted Mean | Weighted SD | Combined N | Number of Studies | Weighted Mean | | Weighted SD | | Combined N | | Number of Studies |
| | | | | | F | M | F | M | F | M | |
| Vision Loss | 6.1 | 1.6 | 254 | 4 | 6.4 | 5.6 | 1.3 | 1.3 | 54 | 63 | 2 |
| Behavioural changes | 8.5 | 3.9 | 194 | 2 | | | | | | | |
| Cognitive decline | 9.3 | 3.1 | 219 | 3 | | | | | | | |
| Epileptic seizures | 10.2 | 3.0 | 243 | 4 | 9.6 | 10.2 | 2.5 | 2.7 | 55 | 63 | 2 |
| Sleep disturbance | 11 | 6.1 | 111 | 1 | | | | | | | |
| Complete blindness | 11.4 | 3.6 | 171 | 3 | | | | | | | |
| Motor decline | 11.0 | 3.8 | 108 | 2 | | | | | | | |
| Speech and language impairment | 12.7 | 4.8 | 136 | 2 | | | | | | | |
| Parkinsonian gait | 14.1 | 2.5 | 111 | 1 | | | | | | | |
| Cardiac manifestations | 17.8 | 4.4 | 45 | 2 | | | | | | | |
| Loss of independent walking | 19.5 | 3.2 | 70 | 3 | | | | | | | |
| Feeding difficulties | 22.0 | 1.6 | 35 | 1 | | | | | | | |
| Death | 22.4 | 4.4 | 95 | 2 | | | | | | | |

**SD**, standard deviation; **M**, male; **F,** female
